# Combinatorial analysis of phenotypic and clinical risk factors associated with hospitalized COVID-19 patients

**DOI:** 10.1101/2021.02.08.21250899

**Authors:** Sayoni Das, Matthew Pearson, Krystyna Taylor, Veronique Bouchet, Gert Lykke Møller, Taryn O. Hall, Mark Strivens, Kathy T. H. Tzeng, Steve Gardner

## Abstract

Characterization of the risk factors associated with variability in the clinical outcomes of COVID-19 is important. Our previous study using genomic data identified a potential role of calcium and lipid homeostasis in severe COVID-19. This study aimed to identify similar combinations of features (disease signatures) associated with severe disease in a separate patient population with purely clinical and phenotypic data.

The PrecisionLife combinatorial analytics platform was used to analyze features derived from de-identified health records in the UnitedHealth Group COVID-19 Data Suite. The platform identified and analyzed 836 disease signatures in two cohorts associated with increased risk of COVID-19 hospitalization. Cohort 1 was formed of cases hospitalized with COVID-19 and a set of controls who developed mild symptoms. Cohort 2 included Cohort 1 individuals for whom additional laboratory test data was available.

We found several disease signatures where lower levels of lipids were found co-occurring with lower levels of serum calcium and leukocytes. Many of the low lipid signatures were independent of statin use and 50% of cases with hypocalcemia signatures were reported with vitamin D deficiency. These signatures may be attributed to similar mechanisms linking calcium and lipid signaling where changes in cellular lipid levels during inflammation and infection affect calcium signaling in host cells.

This study and our previous genomics analysis demonstrate that combinatorial analysis can identify disease signatures associated with the risk of developing severe COVID-19 separately from genomic or clinical data in different populations. Both studies suggest associations between calcium and lipid signalling in severe COVID-19.

## 2 Introduction

The Coronavirus disease 2019 (COVID-19) outbreak caused by the severe acute respiratory syndrome coronavirus 2 (SARS-CoV-2) has been declared a pandemic that has resulted in significant mortality, major social and economic disruption worldwide^1^. As of December 2020, there are over 64 million confirmed cases globally, resulting in more than 1.4 million deaths worldwide^2^. The uncertainty surrounding the progression, management and outcomes of COVID-19 has made it particularly challenging for healthcare systems. Studies have suggested that approximately 80% of COVID-19 positive patients present with mild symptoms or are asymptomatic, and that around 20% of the patients develop a more severe response within 7–14 days from onset of symptoms that may lead to hospitalization and in some cases (2.3%), death^3,4,5,6^.

The risk of developing severe COVID-19 is known to be higher in people who are older, male and have underlying health conditions such as hypertension, cardiovascular disease, diabetes, obesity, chronic respiratory diseases and cancer^5,6^. Approximately 22% of the global population have at least one co-morbidity that puts them at increased risk of severe COVID-19 if exposed to the virus^7^. Ethnicity and socio-economic deprivation have also been associated with severe illness^8^.

SARS-CoV-2 binds to the host cell receptor through the angiotensin-converting enzyme-2 (ACE2)^9^. and starts replicating rapidly inside the host cells, which can trigger a hyperimmune response in some patients^10^. This may be due to generation of pro-inflammatory cytokines and chemokines called a cytokine storm that can cause acute respiratory distress syndrome (ARDS) in the lung and multi-organ failure^11,12^. Other studies have suggested that binding of SARS-CoV-2 increases the levels of ACE2 in lung cells that results in elevated levels of bradykinin^13^ (bradykinin storm) leading to vascular leakage, hypotension and pulmonary edema^14^. These are manifested in COVID-19 patients with pneumonia and respiratory failure. Bradykinin’s role in regulation of clotting may be one mechanism for the extra-pulmonary manifestations such as thromboembolic complications, cardiac events, acute renal and hepatic injury^15,16^. Other symptoms such as neurological complications and gastrointestinal and endocrine symptoms have also been reported^15,17^. Recent evidence suggests that some patients with COVID-19 can also develop long-term complications or experience prolonged symptoms^18,19^.

Early identification and characterization of the risk factors associated with varying clinical outcomes of severely ill COVID-19 patients are crucial for accurate clinical stratification and development of effective management and targeted therapeutic strategies. A previous case-control study using genomic data^20^ identified 68 severe COVID-19 risk-associated protein-coding genes in a population of hospitalized COVID-19 patients in UK Biobank^21,22^. Nine of these were previously linked to differential response to SARS-CoV-2 infection. Several of these genes related to key biological pathways associated with the development of severe COVID-19 and associated symptoms including cytokine production cascades, endothelial cell dysfunction, lipid droplets, calcium signaling, and viral susceptibility factors^20^.

In this study, we identified and assessed the phenotypic and clinical risk factors associated with hospitalized COVID-19 patients in the UnitedHealth Group (UHG) COVID-19 Data Suite using a similar combinatorial analysis approach. Using laboratory test data available for the UHG cohort, we investigated potential correlations with the genomic analysis findings and hypotheses from our previous UK Biobank COVID-19 study^20^, including the potential association of calcium signaling and lipid dysregulation with severe clinical outcomes in COVID-19 patients.

## 3 Method

### Cohort Generation

We used de-identified records of Medicare Advantage and commercially insured members with COVID-19 tests results in the UnitedHealth Group (UHG) COVID-19 Data Suite accessed through the UHG Clinical Discovery Portal for this study. The UHG COVID-19 Data Suite contains longitudinal health information on individuals representing diverse ethnicities, age groups and geographical regions across the United States. The information includes data on COVID-19 test results, in-patient admission data for hospitalized individuals, medical and pharmacy claims, general diagnostic information, demographic data and information on healthcare insurance plans.

We performed case-control studies on two cohorts to identify combinatorial disease signatures associated with the risk of hospitalization for COVID-19 positive patients. Cohort 1, consisting of 9,493 individuals (3,183 cases, 6,310 controls), was generated from the UHG COVID-19 Data Suite (as of August 2020). This contained 3,183 cases who had been hospitalized as a result of developing severe COVID-19 (based on primary diagnosis records) and 6,310 mild controls who had tested positive for COVID-19 but not been hospitalized (**Supplementary Table 1**). Patients who were enrolled in the Medicare Special Needs Plan (SNP) were excluded to reduce any confounding factors associated with these patients, who are often above 65 years old and diagnosed with severe/disabling chronic conditions that increase their risk of hospitalization. Patients without linked clinical data since 2019 were also excluded.

To investigate the potential role of calcium and lipid homeostasis in COVID-19 patients with severe clinical outcomes, we selected five laboratory analytes that were relevant for this hypothesis and had good coverage in Cohort 1. These included serum calcium, low-density cholesterol (LDL), high-density cholesterol (HDL), triglycerides and leukocyte count. A sub cohort, Cohort 2, consisting of 1,581 patients (581 cases and 1,000 controls) was generated for the individuals with laboratory test results for these five analytes.

### Feature generation

The clinical, claims and pharmacy data were converted to categorical features for the study. Flags based on the co-morbidity indices were used, where available. ICD10 diagnosis codes from medical claims that were not included within the co-morbidity flags, and classes of medications listed in pharmacy claims were converted to binary features to reflect incidence in patients since 2019. Quantitative features such as age, laboratory test results and risk scores were also converted to binary features, where values below 0.5 standard deviations (SD) from the mean were assigned the value 0 and those above 0.5 SD were categorized as 1. Any values within ± 0.5 SD were considered missing. Features that were associated with at least one patient (value=1) in each cohort were included in the analyses.

The clinical and phenotypic data available for all individuals in Cohort 1 generated 1,339 binary features per patient (**Supplementary Table 2**). An additional, five laboratory analyte features were added for Cohort 2.

### Combinatorial Analysis

We used the PrecisionLife’s combinatorial multi-omics platform to identify combinations of clinical and phenotypic features for patients from the two cohorts. This analysis allows hypothesis-free, untrained detection of high order, disease associated combinations of features (typically three to ten features in combination known as ‘disease signatures’) that together are strongly associated with a specific disease diagnosis or other clinical phenotype such as fast disease progression or therapy response. This approach has been validated in multiple disease populations^23,24^. Terminology and examples for the mining and analysis process are given in **Supplementary Figure 1**.

The PrecisionLife platform generated statistically significant disease signatures containing up to five features, using a False Discovery Rate (FDR) of 1% and 2,500 cycles of fully random permutations for each dataset. Each analysis took less than an hour to complete, running on a 2 CPU, 4 GPU cloud compute server. These were mapped to the cases in which they were found, and in-patient clinical data were used to generate a patient profile for each combinatorial disease signature. The diseases signatures for each cohort were clustered by the patients in whom they co-occur, generating a network of clinical features associated with the hospitalized patient population.

## 4 Results

### Cohort characteristics

Cohort 1 patients (3,183 cases) had a 19.1% (607 cases) mortality rate, while 51.3% (1,548 cases) were released from care and 29.6% (915 cases) were transferred to other healthcare facilities. Within Cohort 1, 51.3% were female, and 66.7% were Caucasian with a median age of 75 (**Table 1, Supplementary Figure 2**). 54% of the hospitalized patients had at least one of the comorbidities previously linked with higher risk for COVID-19 severe response. Hypertension (52.1%) was the most common co-morbidity, followed by cardiovascular disease (38%), diabetes (31.5%), chronic lung disease (25.9%) and dementia (13.9%) (**Table 1**). The most common COVID-19 related diagnoses reported in hospital admissions data for cases were pneumonia (43%), followed by respiratory failure (18.3%) and septicemia (7.3%) (**Supplementary Figure 3**).

**Table 1.** Cohort characteristics for the hospitalization risk studies. *p-values* were calculated to assess the association of each feature in the two COVID-19 hospitalized risk cohorts using two-sided Fisher’s exact tests for categorical data and Mann-Whitney U tests for continuous data.

|  | Cohort 1 (n = 9,493) |  |  | Cohort 2 (n = 1,581) |  |  |
| --- | --- | --- | --- | --- | --- | --- |
|  | Hospitalized patients | Non-Hospitalized patients | Two-sided *p-value* | Hospitalized patients | Non-Hospitalized patients | Two-sided *p-value* |
| **COVID-19 positive** | 3,183 | 6,310 | N/A | 581 | 1,000 | N/A |
| **Males (%)** | 1,549 (48.7%) | 2,758 (43.7%) | 5.0e-06 | 295 (50.1%) | 348 (34.8%) | 6.5e-10 |
| **Median Age [Range]** | 75 [29-89] | 72 [15-89] | <2.2e-16 | 74 [31-89] | 71.5 [33-89] | 2.2e-14 |
| **Caucasian\* (%)** | 1,632 (66.7%) | 3,693 (66.8%) | 0.938 | 298 (57.5%) | 528 (59.5%) | 0.465 |
| **Mortality (%)** | 607 (19.10%) | N/A | N/A | 100 (17.2%) | N/A | N/A |
| **Hypertension (%)** | 1,657 (52.1%) | 3,864 (61.2%) | <2.2e-16 | 431 (58.7%) | 672 (67.2%) | 3.7e-03 |
| **Diabetes (%)** | 1,109 (34.8%) | 2,114 (33.5%) | 0.198 | 277 (47.7%) | 427 (42.7%) | 5.8e-02 |
| **Cardiovascular (%)** | 1,210 (38.0%) | 1,468 (23.3%) | <2.2e-16 | 204 (35.1%) | 267 (26.7%) | 2.2e-03 |
| **Dementia (%)** | 443 (13.9%) | 236 (3.7%) | <2.2e-16 | 27 (4.6%) | 10 (1.0%) | 7.2e-06 |
| **Chronic lung disease (%)** | 832 (25.9%) | 1,198 (20.0%) | 2.4e-15 | 135 (23.2%) | 188 (18.9%) | 3.8e-02 |

\* The percentage of Caucasians was calculated as a fraction of those individuals for whom ethnicity information was available (approximately 85% of all records).

### Combinatorial disease signatures capture phenotypic and clinical risk factors for severe COVID-19

Combinatorial analysis identified 1,147 combinations of clinical and phenotypic features (disease signatures) that were highly associated with hospitalized patients in Cohort 1 and 32,242 combinations in Cohort 2 (**Supplementary Table 3, Supplementary Figure 4**). Higher number of disease signatures were reported for Cohort 2. This is likely due to relatively higher prevalence of the same clinical features among Cohort 2 individuals as compared to Cohort 1.

The disease signatures were filtered to exclude those that had any features indicating absence of a disease diagnosis, symptom or medication use, as these are likely to be generated as a result of incompleteness of the claims and pharmacy data rather than as a true disease association. Additionally, disease signatures that were found in fewer than 20 cases were also excluded. After filtering, 255 disease signatures in Cohort 1 and 531 disease signatures in Cohort 2 were used for further analysis.

All features in the disease signatures identified for each study were scored using a Random Forest (RF) algorithm based inside a 5-fold cross-validation framework to evaluate the accuracy with which a feature (e.g. a laboratory analyte value) predicts the observed case:control split (minimizing Gini impurity). 166 features in Cohort 1 and 41 features in Cohort 2 were identified as critical features as shown in **Supplementary Figure 5**. Many of these included diagnoses and symptoms associated with severe COVID-19 such as respiratory failure, pneumonia, acute renal failure and septicemia because of their low incidence in controls.

We found that the combinatorial disease signatures capture clinical features associated with response to severe COVID-19 illness (**Figure 1, Figure 2**) These features include pneumonia and respiratory failure, which are frequently reported among hospitalized patients, and risk factors that increase the probability of developing severe response such as diabetes, hypertension and cardiovascular disease. Phenotypes related to the risk-associated comorbidities such as elevated glucose levels or blood pressure and common medications prescribed for them (e.g. insulin, statins and dihydropyridines) were also commonly found. Many low frequency features (<10% among hospitalized patients) such as ARDS^11^, pneumothorax^25^, hematuria^26^, encephalopathy^17^, pericarditis^27^ and thrombosis^15^ were frequently found in disease signatures in combination with other features. Some disease signatures also captured clinical features related to increased frailty such as senility or high risk of hospital-readmission, whilst other features reflect conditions that are associated with prolonged hospital stay such as pressure ulcers and secondary bacterial infections.

**Figure 1.**
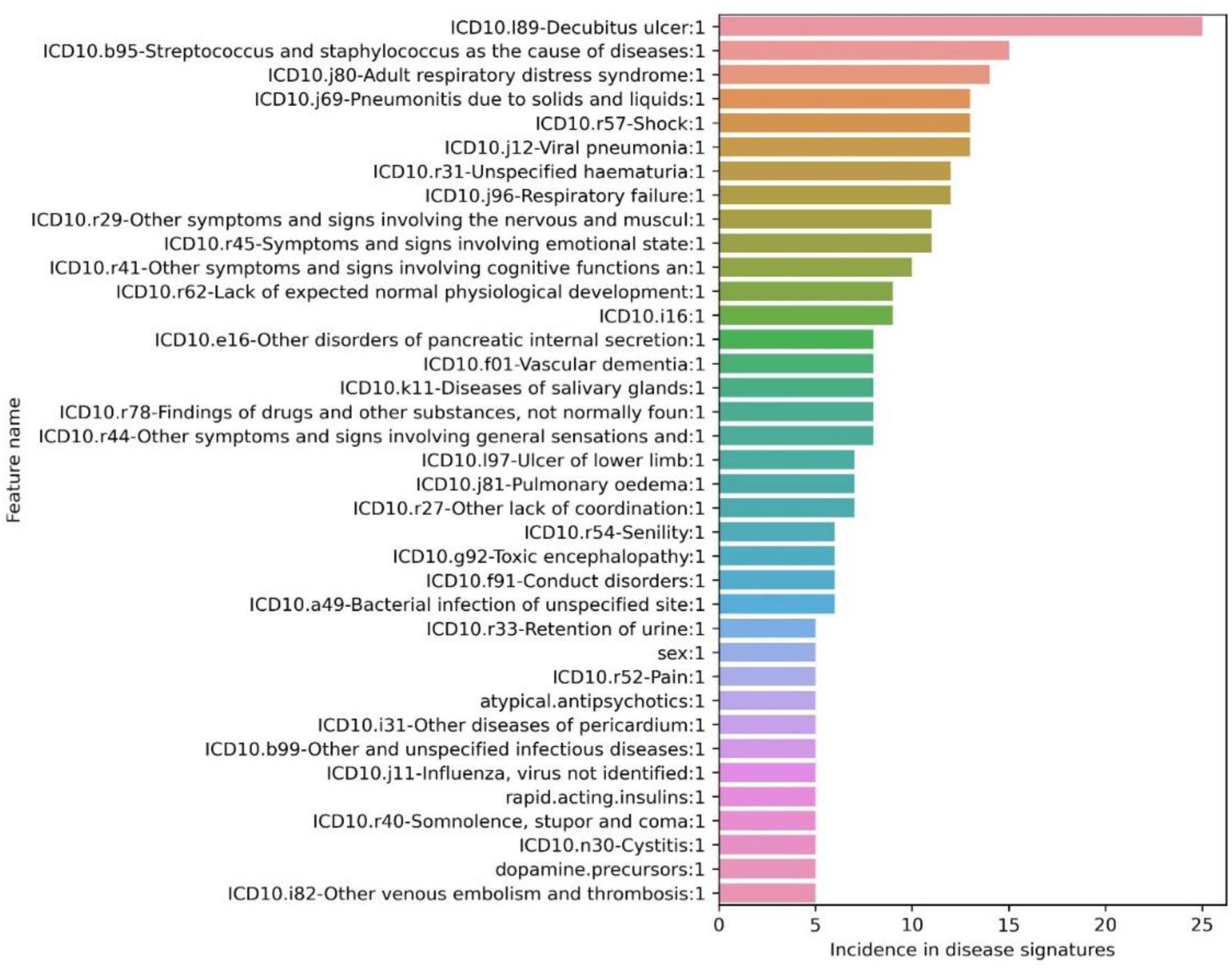
Phenotypic and clinical features that were most frequently reported in 255 filtered disease signatures in Cohort 1 associated with increased risk of hospitalization with severe COVID-19.

**Figure 2.**
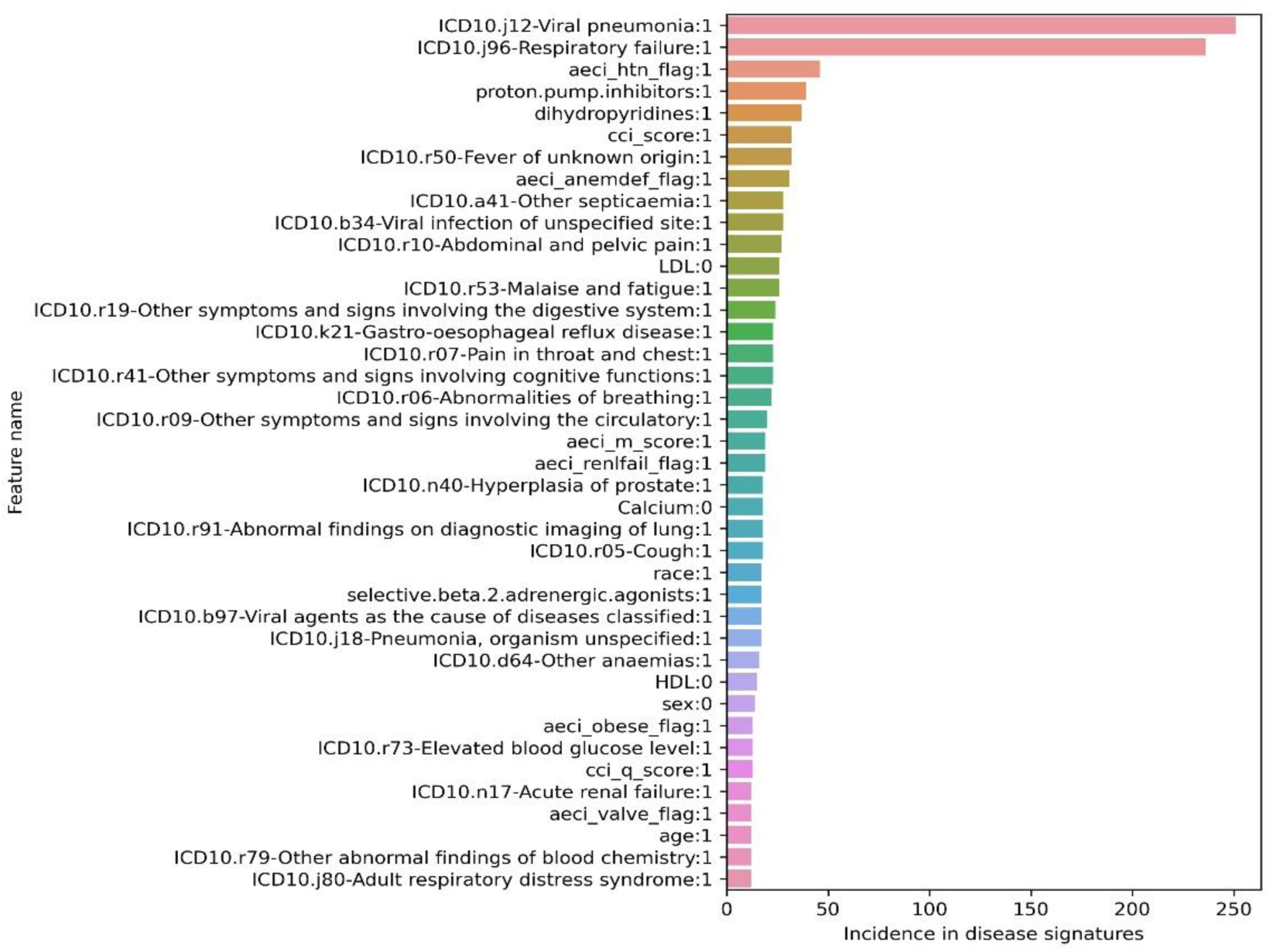
Phenotypic and clinical features that were most frequently reported in 581 filtered disease signatures in Cohort 2 (sub cohort of Cohort 1 with additional laboratory test results) associated with increased risk of hospitalization with severe COVID-19. Features associated with hypocalcemia (Calcium:0) and hypolipidemia (LDL:0, HDL:0) were reported in multiple disease signatures.

Networks generated by clustering disease signatures in the two cohorts highlighted the heterogeneity of clinical features observed in severe COVID-19. Such clustering enables identification of disease signatures that co-occur in patient sub-groups who are likely to have similar symptoms, underlying conditions or clinical outcomes. For example, hospitalized patients who developed ARDS were likely to be influenced by the features nearest to ARDS in the network such as older age, development of pneumonia, pulmonary hemorrhage, sepsis and high mortality (**Figure 3, Supplementary Figure 6**).

**Figure 3.**
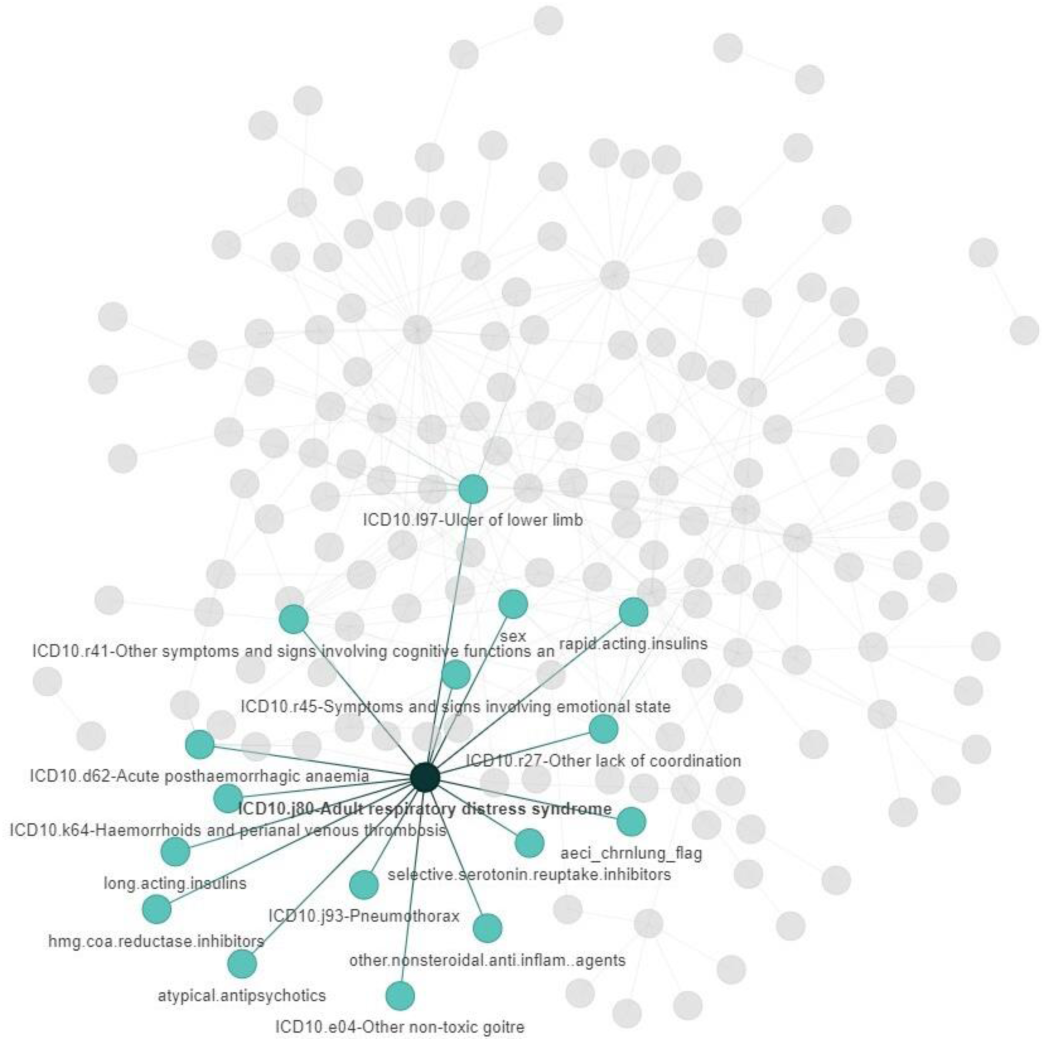
Network architecture of filtered (n=255) disease signatures associated with hospitalized COVID-19 patients in Cohort 1 generated by the PrecisionLife platform where each circle represents a feature and edges represent co-association in patients. The colored nodes and edges represent the disease signatures of patients who developed ARDS (shown in darker shade) in Cohort 1. The co-associated features are shown in lighter shade.

### Disease signatures associated with lower levels of serum calcium and lipids

In Cohort 2, features from five blood analytes (calcium, LDL, HDL, triglycerides and leukocyte count) were available for patients. Hospitalized patients with severe COVID-19 were observed to be more likely to have lower serum calcium levels (<9.26 mg/dl), lower LDL levels (<78.23 mg/dl), lower HDL levels (<44.35 mg/dl) and higher levels of triglycerides (>206.20 mg/dl) when compared against the patients with mild disease (**Supplementary Table 4**). Both low and high levels of blood leukocyte count were observed in patients with severe COVID-19.

In Cohort 2 the PrecisionLife platform identified 18 disease signatures in 80 hospitalized patients with serum calcium values lower than 9.26 mg/dl (**Supplementary Figure 7**). Out of these, only four signatures were co-associated with the use of the dihydropyridines (calcium channel blockers) and proton-pump inhibitors which may have an effect on calcium homeostasis^28,29^. The hypocalcemia disease signatures were associated with COVID-19 symptoms such as pneumonia and respiratory failure, and comorbidities including diabetes, hypertension and anemia. Two calcium disease signatures were found in 34 patients (42.5%), co-occurring with high mortality and hospital re-admission risk scores, which suggests that these patients had multiple underlying conditions. Another calcium disease signature in 33 (41.3%) patients was associated with low serum levels of HDL and pneumonia.

We also identified 45 disease signatures in 188 (32.4%) severe COVID-19 patients that were associated with comparatively low serum lipid (LDL, HDL or triglyceride) levels (**Supplementary Figures 8-10**). Comorbidities such as hypertension, obesity and cerebrovascular disease were found in these hypolipidemia signatures, which are which are not commonly co-associated in patients. We investigated whether the reduced lipid levels observed in these patients were caused by the use of statins. None of the disease signatures were associated with the feature indicating statin use by all associated cases. We found 12 hypolipidemia signatures where less than 10% of the patients were associated with any prescription records for statins within 90 days of the laboratory test result date, suggesting that these signatures were independent of statin use. Thus, dyslipidemia observed in many severe COVID-19 patients in Cohort 2 is not likely to represent an artefact of other comorbidities or medication use, but a consequential host response to SARS-CoV-2 infection which has been reported in many recent studies^30,31,32^.

Mortality in the patients with either calcium or lipid disease signatures was not found to be significantly different. We were able to identify 15 disease signatures with lower levels of calcium and one signature with lower levels of cholesterol in this sub cohort that were associated with at least 10 patients. The identification of calcium and lipid disease signatures in this sub cohort strongly suggests that they reflect biochemical characteristics of patients with severe host response to COVID-19.

## 5 Discussion

Pulmonary manifestations of COVID-19 such as respiratory failure and pneumonia were the most common symptoms in the two cohorts that were also prevalent in the combinatorial disease signatures identified by the PrecisionLife platform (**Supplementary Figure 3, Supplementary Figure 5**). Comorbidities such as hypertension, cardiovascular disease, chronic respiratory disease and diabetes known to be associated with COVID-19 risk from other studies^3,4,5^, including our previous genetic study^19^ in UK Biobank, were observed in hospitalized patients. These comorbidities co-occur with different COVID-19 symptoms, complications, medication use and laboratory analyte values. This analysis enables us to gain useful insights into the likely associations between these clinical and phenotypic features that can improve clinical management of patients.

A wide variety of severe COVID-19 manifestations such as ARDS, sepsis, pericarditis and thrombosis were observed in the disease signatures representing patient sub-groups^3,15,25,26,27^. This correlates with our previous genomic analysis on the UK Biobank COVID-19 cohort, which identified genes associated with some of these complications, including host pathogenic responses, inflammatory cytokine production, modulation of cardiac function and endothelial cell function^20.^

Use of medications such as proton pump inhibitors, dihydropyridines and beta‐adrenergic blockers were observed in seven disease signatures in Cohort 1 and 80 signatures in Cohort 2. Dihydropyridines^33,34^ and beta‐adrenergic blockers^35,36^ have been associated with improved outcomes for COVID-19 patients and suggested as potential treatments, while proton pump inhibitors have been associated with adverse outcomes in several studies^37,38^. The incidence of the medications in the disease signatures could be either due to adverse effects cause by the medication resulting in more severe COVID-19 response or it could reflect the comorbidities in patients for which they are generally prescribed. Using the available data, it was not possible for us to ascertain the specific association of these medications in our study with certainty.

In Cohort 2, all hypocalcemia (n=18) disease signatures and hypolipidemia (n=45) signatures were found to be associated with severe pulmonary manifestations of COVID-19 (**Supplementary Figures 7-10**). There is increasing evidence that calcium and lipid homeostasis play an important role in the viral replication cycle and they have been suggested as biomarkers for increased COVID-19 severity^30,31,32,39^. It has been demonstrated that calcium signaling pathway or calcium-dependent processes in host cells are often perturbed by viral proteins that can bind calcium and/or calcium-binding protein domains, allowing them to modulate the host cellular machinery for viral replication, assembly and release^40,41^. The mechanism of calcium regulation is not fully understood, as some viruses are known to increase intracellular calcium levels while others are known to have a dynamic control based on the phase of infection^42^. However, the SARS-CoV E protein has been shown to form protein-lipid channels that transport calcium ions, activating the NLRP3 inflammasome and increasing systemic inflammation via IL-1β^43^.

Lower lipid levels have been reported in severe COVID-19 patients in many studies with a correlation observed between reduced lipid levels and disease severity^44,45,46^. Many viruses, including SARS-CoV and MERS-CoV, can modulate lipid synthesis and signaling in host cells to divert cellular lipids to viral replication and exocytosis, facilitating invasion of other host cells^47,48^. It has been suggested that the decrease in cellular cholesterol levels following SARS-CoV-2 infection leads to disruption of the signaling hub for inflammation and cholesterol metabolism, resulting in the dysregulation of cholesterol biosynthesis, inflammatory cytokine release and vascular homeostasis^49,50^.

Regulation of cholesterol biosynthesis has been shown to be associated with six genes identified by a genome-scale CRISPR knockout screen that reduced SARS-Cov-2 infection in human alveolar basal epithelial carcinoma cells^51^. The study also demonstrated that use of dihydropyridines results in increased resistance to SARS-Cov-2 infection^51^. Another study hypothesized that elevated unsaturated fatty acids in SARS-CoV-2 infected host cells bind calcium, resulting in hypocalcemia and triggering the production of pro-inflammatory mediators and cytokine storm induction^52,53^.

We found seven disease signatures in this study where lower levels of LDL were found co-occurring with lower levels of serum calcium, leukocyte count or HDL. These signatures may be attributed to similar mechanisms linking calcium and lipid signaling where changes in cellular lipid levels during inflammation and infection^54^ affects calcium signaling in host cells^55,56,57^.

Retrospective analysis of the clinical histories of the hospitalized patients with lower calcium and lipid signatures were performed to identify whether the laboratory analyte values may be affected by other medical conditions. We found that 50% of cases represented by diseases signatures featuring lower levels of calcium were reported to have vitamin D deficiency. More than 25% of people above the age of 65 are vitamin D deficient, with vitamin D playing a key role in calcium homeostasis^58^. This suggests that the changes in calcium levels in patients in this study may be linked to vitamin D deficiency in severe COVID-19^58,59^, which has also been associated with severe illness and which was found in eight disease signatures in Cohort 2. Our previous analysis on the UK Biobank COVID-19 cohort^20^ identified 16 calcium-binding/signaling genes and six genes relating to lipid droplet biology and correlated with serum lipid levels and coronary artery disease. In conjunction with the findings of this study, this adds further support to the role of calcium and lipid signaling in relation to viral pathogenesis and severe host response to COVID-19. To fully understand the role of calcium and lipid homeostasis in COVID-19, analysis of patient datasets that combine genetic, clinical and hospital laboratory test data will be necessary.

### Limitations of the Study

This study was limited by the completeness of data for features relevant to analyzing differential host response to COVID-19. Information on the onset of disease or symptoms, clinical phase of disease, viral load, oxygen saturation, breathing rate, body mass index and physiological measurements or biomarker levels during hospitalization was not consistently available. We used hospitalization status associated with primary diagnosis of COVID-19 to as surrogate for severe COVID-19 patients. Mortality and diagnoses linked to clinical progression of COVID-19 were used to estimate relative severity of disease among hospitalized patients.

The comorbidities, diagnoses, medications and laboratory test results were derived from medical claims, pharmacy claims and in-patient admission records. Since claims data are generated for reimbursement and administrative purposes rather than scientific research, the records may be missing information and there is potential for variability in their collection. Data sparsity of the available patient records was reflected in the low penetrance of many disease signatures. As more patient data becomes available, the disease signatures will become more predictive, enabling higher resolution patient stratification.

## 6 Conclusion

The PrecisionLife platform identified and analyzed 836 combinatorial disease signatures in two COVID-19 cohorts (Cohort 1=255, Cohort 2=531) associated with increased risk of hospitalization from COVID-19. These disease signatures were found to capture different symptomatic presentations of COVID-19, complications arising from the clinical progression of the disease, and underlying disease conditions that could be either associated with severe host response to COVID-19 or were indicative of conditions associated with older age or frailty.

In the second cohort, we found 45 disease signatures that were associated with lower levels of serum calcium, LDL, HDL and triglycerides in 188 (32.35%) hospitalized patients. This suggests that lower levels of calcium and cholesterol are biochemical characteristics associated with severe COVID-19 patients, which may also add further support to the role of calcium signaling and lipid dysregulation in SARS-CoV-2 pathogenesis. This also validates our findings from our previous genomics study^20^ on severe COVID-19 patients in UK Biobank^21^ where we identified 16 risk-associated genes that had calcium-binding domains or were involved in calcium signaling and six genes linked to lipid droplet biology associated with serum lipid levels.

This study along with our previous genomic study^20^ demonstrates that a combinatorial analysis approach is able to identify related groups of clinical and phenotypic features from both genomic and phenotypic data that are associated with risk of developing severe forms of COVID-19. This enables us to gain useful insights into the likely associations between the features of interest that could improve clinical management of patients. With the availability of more data, the combinatorial output of the analytical platform would be greatly enhanced.

This analysis also validates the association of calcium and lipid homeostasis with severe COVID-19 reported by our previous study, using real-world data in an independent cohort. This study will extend these analyses in future to larger patient datasets that have both genetic and phenotypic data to fully ascertain the differences between mild and severe host responses to COVID-19 and the mechanism of calcium and lipid signaling in SARS-Cov-2 pathogenesis.

## Data Availability

The data analyzed in this study was obtained from UnitedHealth Group Clinical Discovery Portal. The data are proprietary and are not available for public use but, under certain conditions, may be made available to editors and their approved auditors under a data-use agreement to confirm the findings of the current study. Further inquiries can be directed to Scott Schneweis.

## 8 Acknowledgements

We would like to acknowledge the UnitedHealth Group for providing us access to the COVID-19 Data Suite through the UHG Clinical Discovery Portal and the patients who provided their data. Special thanks to Megan Jarvis, Kae Tanudtanud, Yinglong Guo, Elena Fultz, Aditya Yellepeddi and Teodi Enrik Racho from the UnitedHealth Group and the rest of the PrecisionLife team for their technical assistance and helpful discussions.

## 9 Conflict of Interest

All authors are employees of their respective companies.

## 10 Author Contributions

SG conceived and supervised the study. MP and SD performed the studies and analyzed the data. SD wrote the manuscript. KT contributed to the study design, analysis of disease signatures and manuscript. VB and MAS contributed to the study design and manuscript. GLM developed the core technology in PrecisionLife’s platform. TOH and KTZ contributed to the study design and coordinated access to the COVID-19 Data Suite through the UHG Clinical Discovery Portal. All authors contributed to the study and approved the final version of the manuscript.

## 11 Funding

No external funding was used for this research.

## 12 Supplementary Material

The Supplementary Material for this article can be found online.

## Notes

### Author Declarations

The study was reviewed by the Office of Human Research Affairs (OHRA) of the UnitedHealth Group. The Federalwide Assurance number of OHRA is FWA00028881 and OHRP Registration number is IORG0010356. Ethical approval for this study was waived by OHRA with the following decision: The research was determined to be exempt category 4 given that the research involves secondary use of existing data (de-identified health records), there will be no interaction with participants and participant data will not be re-identified.

### Summary of Updates

The Author Affiliations were corrected.

## References

1. Cucinotta D, Vanelli M. WHO declares COVID-19 a pandemic. Acta Bio Medica: Atenei Parmensis. 2020 91(1):157. doi:10.23750/abm.v91i1.9397

2. Dong E, Du H, Gardner L. An interactive web-based dashboard to track COVID-19 in real time. The Lancet infectious diseases. 2020 20(5):533–4. doi:10.1016/S1473-3099(20)30120-1

3. Verity R, Okell LC, Dorigatti I, Winskill P, Whittaker C, Imai N, Cuomo-Dannenburg G, Thompson H, Walker PG, Fu H, Dighe A. Estimates of the severity of coronavirus disease 2019: a model-based analysis. The Lancet infectious diseases. 2020 20:669–677. doi:10.1016/S1473-3099(20)30243-7

4. Wu Z, McGoogan JM. Characteristics of and important lessons from the coronavirus disease 2019 (COVID-19) outbreak in China: summary of a report of 72 314 cases from the Chinese Center for Disease Control and Prevention. Jama. 2020 323(13):1239–42.

5. Zhou F, Yu T, Du R, Fan G, Liu Y, Liu Z, Xiang J, Wang Y, Song B, Gu X, Guan L. Clinical course and risk factors for mortality of adult inpatients with COVID-19 in Wuhan, China: a retrospective cohort study. The Lancet. 2020 395(10229):1054–1062.

6. Hu B, Guo H, Zhou P, Shi ZL. Characteristics of SARS-CoV-2 and COVID-19. Nature Reviews Microbiology. 2020.

7. Clark A, Jit M, Warren-Gash C, Guthrie B, Wang HH, Mercer SW, Sanderson C, McKee M, Troeger C, Ong KL, Checchi F. Global, regional, and national estimates of the population at increased risk of severe COVID-19 due to underlying health conditions in 2020: a modelling study. The Lancet Global Health. 2020 8(8):e1003–17.

8. Niedzwiedz CL, O’Donnell CA, Jani BD, Demou E, Ho FK, Celis-Morales C, Nicholl BI, Mair FS, Welsh P, Sattar N, Pell JP. Ethnic and socioeconomic differences in SARS-CoV-2 infection: prospective cohort study using UK Biobank. BMC medicine. 2020 18:1–4.

9. Hoffmann M, Kleine-Weber H, Schroeder S, Krüger N, Herrler T, Erichsen S, Schiergens TS, Herrler G, Wu NH, Nitsche A, Müller MA. SARS-CoV-2 cell entry depends on ACE2 and TMPRSS2 and is blocked by a clinically proven protease inhibitor. Cell. 2020 181(2):271-280.e8.

10. Zhou P, Yang XL, Wang XG, Hu B, Zhang L, Zhang W, Si HR, Zhu Y, Li B, Huang CL, Chen HD. A pneumonia outbreak associated with a new coronavirus of probable bat origin. nature. 2020 579(7798):270–273.

11. Ragab D, Salah Eldin H, Taeimah M, Khattab R, Salem R. The COVID-19 cytokine storm; what we know so far. Frontiers in immunology. 2020 16;11:1446.

12. Coperchini F, Chiovato L, Croce L, Magri F, Rotondi M. The cytokine storm in COVID-19: an overview of the involvement of the chemokine/chemokine-receptor system. Cytokine & Growth Factor Reviews. 2020 53:25–32.

13. Garvin MR, Alvarez C, Miller JI, Prates ET, Walker AM, Amos BK, Mast AE, Justice A, Aronow B, Jacobson D. A mechanistic model and therapeutic interventions for COVID-19 involving a RAS-mediated bradykinin storm. Elife. 2020 9:e59177.

14. Zwaveling S, van Wijk RG, Karim F. Pulmonary edema in COVID-19: Explained by bradykinin?. Journal of Allergy and Clinical Immunology. 2020 146(6):1454–5.

15. Gupta A, Madhavan MV, Sehgal K, Nair N, Mahajan S, Sehrawat TS, Bikdeli B, Ahluwalia N, Ausiello JC, Wan EY, Freedberg DE. Extrapulmonary manifestations of COVID-19. Nature medicine. 2020 26(7):1017–32.

16. Gavriatopoulou M, Korompoki E, Fotiou D, Ntanasis-Stathopoulos I, Psaltopoulou T, Kastritis E, Terpos E, Dimopoulos MA. Organ-specific manifestations of COVID-19 infection. Clinical and experimental medicine. 2020.

17. Garg RK, Paliwal VK, Gupta A. Encephalopathy in patients with COVID‐ 19: a review. Journal of Medical Virology. 2020.

18. Iacobucci G. Long covid: Damage to multiple organs presents in young, low risk patients. BMJ: British Medical Journal (Online). 2020 371.

19. Dennis A, Wamil M, Kapur S, Alberts J, Badley A, Decker GA, Rizza SA, Banerjee R, Banerjee A. Multi-organ impairment in low-risk individuals with long COVID. medrxiv. 2020.

20. Taylor K, Das S, Pearson M, Kozubek J, Pawlowski M, Jensen CE, Skowron Z, Møller GL, Strivens M, Gardner S. Analysis of genetic host response risk factors in severe COVID-19 patients. medRxiv. 2020.

21. Bycroft C, Freeman C, Petkova D, Band G, Elliott LT, Sharp K, Motyer A, Vukcevic D, Delaneau O, O’Connell J, Cortes A. The UK Biobank resource with deep phenotyping and genomic data. Nature. 2018 562(7726):203–9.

22. Armstrong J, Rudkin JK, Allen N, Crook DW, Wilson DJ, Wyllie DH, O’Connell AM. Dynamic linkage of covid-19 test results between public health england’s second generation surveillance system and uk biobank. Microbial genomics. 2020 6(7).

23. Koefoed P, Andreassen OA, Bennike B, Dam H, Djurovic S, Hansen T, Jorgensen MB, Kessing LV, Melle I, Møller GL, Mors O. Combinations of SNPs related to signal transduction in bipolar disorder. PloS one. 2011 6(8):e23812.

24. Taylor K, Das S, Pearson M, Kozubek J, Strivens M, Gardner S. Systematic drug repurposing to enable precision medicine: A case study in breast cancer. Digital Medicine. 2019 5(4):180.

25. Zantah M, Castillo ED, Townsend R, Dikengil F, Criner GJ. Pneumothorax in COVID-19 disease-incidence and clinical characteristics. Respiratory Research. 2020 21(1):1–9.

26. Liu X, Zhang R, He G. Hematological findings in coronavirus disease 2019: indications of progression of disease. Annals of Hematology. 2020.

27. Tung-Chen Y. Acute pericarditis due to COVID-19 infection: An underdiagnosed disease?. Medicina Clinica (English Ed.). 2020 155(1):44.

28. Price D, Radke J, Albertson T. Hypocalcaemia after an occult calcium channel blocker overdose: a case report and literature review. Basic & clinical pharmacology & toxicology. 2014 Feb;114(2):217– 21.

29. Sivakumar J. Proton pump inhibitor-induced hypomagnesaemia and hypocalcaemia: case review. International journal of physiology, pathophysiology and pharmacology. 2016;8(4):169.

30. Sun JK, Zhang WH, Zou L, Liu Y, Li JJ, Kan XH, Dai L, Shi QK, Yuan ST, Yu WK, Xu HY. Serum calcium as a biomarker of clinical severity and prognosis in patients with coronavirus disease 2019. Aging (Albany NY). 2020 12(12):11287.

31. di Filippo L, Formenti AM, Doga M, Frara S, Rovere-Querini P, Bosi E, Carlucci M, Giustina A. Hypocalcemia is a distinctive biochemical feature of hospitalized COVID-19 patients. Endocrine. 2020.

32. Yang C, Ma X, Wu J, Han J, Zheng Z, Duan H, Liu Q, Wu C, Dong Y, Dong L. Low serum calcium and phosphorus and their clinical performance in detecting COVID‐ 19 patients. Journal of medical virology. 2020.

33. Solaimanzadeh I. Nifedipine and Amlodipine Are Associated With Improved Mortality and Decreased Risk for Intubation and Mechanical Ventilation in Elderly Patients Hospitalized for COVID-19. Cureus. 2020 12(5).

34. Zhang L, Sun Y, Zeng HL, Peng Y, Jiang X, Shang WJ, Wu Y, Li S, Zhang YL, Yang L, Chen H. Calcium channel blocker amlodipine besylate is associated with reduced case fatality rate of COVID-19 patients with hypertension. medRxiv. 2020.

35. Vasanthakumar N. Beta‐Adrenergic Blockers as a Potential Treatment for COVID‐ 19 Patients. BioEssays. 2020 42(11):2000094.

36. Vasanthakumar N. Can beta-adrenergic blockers be used in the treatment of COVID-19?. Medical Hypotheses. 2020 142:109809.

37. Lee SW, Ha EK, Yeniova AÖ, Moon SY, Kim SY, Koh HY, Yang JM, Jeong SJ, Moon SJ, Cho JY, Yoo IK. Severe clinical outcomes of COVID-19 associated with proton pump inhibitors: a nationwide cohort study with propensity score matching. Gut. 2020 70(1):76–84.

38. Almario CV, Chey WD, Spiegel BM. Increased risk of COVID-19 among users of proton pump inhibitors. The American Journal of Gastroenterology. 2020 115(10):1707–1715.

39. Wei X, Zeng W, Su J, Wan H, Yu X, Cao X, Tan W, Wang H. Hypolipidemia is associated with the severity of COVID-19. Journal of Clinical Lipidology. 2020.

40. Zhou Y, Frey TK, Yang JJ. Viral calciomics: interplays between Ca2+ and virus. Cell calcium. 2009 46(1):1–7.

41. Chen X, Cao R, Zhong W. Host Calcium Channels and Pumps in Viral Infections. Cells. 2020 9(1):94.

42. Moreno-Altamirano MM, Kolstoe SE, Sánchez-García FJ. Virus control of cell metabolism for replication and evasion of host immune responses. Frontiers in cellular and infection microbiology. 2019 9:95.

43. Nieto-Torres JL, Verdiá->Báguena C, Jimenez-Guardeño JM, Regla-Nava JA, Castaño-Rodriguez C, Fernandez-Delgado R, Torres J, Aguilella VM, Enjuanes L. Severe acute respiratory syndrome coronavirus E protein transports calcium ions and activates the NLRP3 inflammasome. Virology. 2015 485:330–9.

44. Wang G, Zhang Q, Zhao X, Dong H, Wu C, Wu F, Yu B, Lv J, Zhang S, Wu G, Wu S. Low high-density lipoprotein level is correlated with the severity of COVID-19 patients: an observational study. Lipids in health and disease. 2020 19(1):1–7.

45. Fan J, Wang H, Ye G, Cao X, Xu X, Tan W, Zhang Y. Low-density lipoprotein is a potential predictor of poor prognosis in patients with coronavirus disease 2019. Metabolism. 2020 154243.

46. Hu X, Chen D, Wu L, He G, Ye W. Declined serum high density lipoprotein cholesterol is associated with the severity of COVID-19 infection. Clinica Chimica Acta. 2020 510:105–10.

47. Abu-Farha M, Thanaraj TA, Qaddoumi MG, Hashem A, Abubaker J, Al-Mulla F. The Role of Lipid Metabolism in COVID-19 Virus Infection and as a Drug Target. International Journal of Molecular Sciences. 2020 21(10):3544.

48. Yan B, Chu H, Yang D, Sze KH, Lai PM, Yuan S, Shuai H, Wang Y, Kao RY, Chan JF, Yuen KY. Characterization of the lipidomic profile of human coronavirus-infected cells: Implications for lipid metabolism remodeling upon coronavirus replication. Viruses. 2019 11(1):73.

49. Guo C, Chi Z, Jiang D, Xu T, Yu W, Wang Z, Chen S, Zhang L, Liu Q, Guo X, Zhang X. Cholesterol homeostatic regulator SCAP-SREBP2 integrates NLRP3 inflammasome activation and cholesterol biosynthetic signaling in macrophages. Immunity. 2018 49(5):842–56.

50. Lee W, Ahn JH, Park HH, Kim HN, Kim H, Yoo Y, Shin H, Hong KS, Jang JG, Park CG, Choi EY. COVID-19-activated SREBP2 disturbs cholesterol biosynthesis and leads to cytokine storm. Signal transduction and targeted therapy. 2020 5(1):1–1.

51. Daniloski Z, Jordan TX, Wessels HH, Hoagland DA, Kasela S, Legut M, Maniatis S, Mimitou EP, Lu L, Geller E, Danziger O. Identification of required host factors for SARS-CoV-2 infection in human cells. Cell. 2020 S0092-8674(20)31394-5.

52. Thomas T, Stefanoni D, Reisz JA, Nemkov T, Bertolone L, Francis RO, Hudson KE, Zimring JC, Hansen KC, Hod EA, Spitalnik SL. COVID-19 infection results in alterations of the kynurenine pathway and fatty acid metabolism that correlate with IL-6 levels and renal status. medRxiv. 2020.

53. Singh VP, Khatua B, El-Kurdi B, Rood C. Mechanistic basis and therapeutic relevance of hypocalcemia during severe COVID-19 infection. Endocrine. 2020 70(3):461–2.

54. Feingold KR, Anawalt B, Boyce A, Chrousos G, Dungan K, Grossman A, Hershman JM, Kaltsas G, Koch C, Kopp P, Korbonits M. The Effect of Inflammation and Infection on Lipids and Lipoproteins--Endotext.

55. Greineisen WE, Speck M, Shimoda LM, Sung C, Phan N, Maaetoft-Udsen K, Stokes AJ, Turner H. Lipid body accumulation alters calcium signaling dynamics in immune cells. Cell calcium. 2014 56(3):169–80.

56. Mori S, Ito H, Yamamoto K. Effects of calcium antagonists on low density lipoprotein metabolism in human arterial smooth muscle cells. The Tohoku Journal of Experimental Medicine. 1988;154(4):329–33.

57. Ranganathan S, Harmony JA, Jackson RL. Effect of Ca2+ blocking agents on the metabolism of low density lipoproteins in human skin fibroblasts. Biochemical and Biophysical Research Communications. 1982 107(1):217–24.

58. Jain A, Chaurasia R, Sengar NS, Singh M, Mahor S, Narain S. Analysis of vitamin D level among asymptomatic and critically ill COVID-19 patients and its correlation with inflammatory markers. Scientific reports. 2020 10(1):1–8.

59. Vyas N, Kurian SJ, Bagchi D, Manu MK, Saravu K, Unnikrishnan MK, Mukhopadhyay C, Rao M, Miraj SS. Vitamin D in prevention and treatment of COVID-19: current perspective and future prospects. Journal of the American College of Nutrition. 2020.

