## Supplementary Material for "Combinatorial analysis of phenotypic and clinical risk factors associated with hospitalized COVID-19 patients"

### 1 Mining Terminology and Example

The overall process of mining, validation and scoring is shown below.

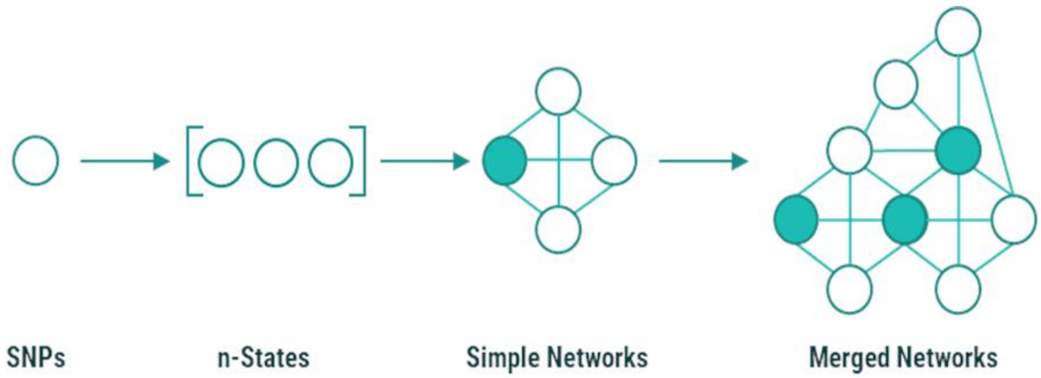


**Supplementary Figure 1.** Stages of the mining, scoring and analysis process in PrecisionLife's combinatorial analytics platform.

### 2 Supplementary Data

**Supplementary Table 1.** COVID-19 positive patient records available in the UnitedHealth Group (UHG) COVID-19 Data Suite.

| | Hospitalized patients | Non-hospitalized patients |
|---|---|---|
| COVID-19 positive patients | 37,027 | 219,271 |
| COVID-19 patients with any clinical data since 2019 | 19,244 | 28,408 |
| Cohort 1 - COVID-19 patients with clinical data who were not enrolled in Medicare Special Needs Plan | 3,183 | 6,310 |
| Cohort 2 - Cohort 1 patients with additional laboratory data for 5 lab analytes | 581 | 1,000 |

**Supplementary Table 2.** Features used for Hospitalization risk studies.

| Feature group | Data source | Description | Feature count |
|---|---|---|---|
| Sex | Member information | 1 indicates Male, 0 indicates Female, 3 indicates missing data | 1 |
| Race | Member information | 1 indicates non-Caucasian, 0 indicates Caucasian and 3 indicates missing data. | 1 |
| Co-morbidity flags based on Co-morbidity Index | Co-morbidity scores table derived by UHG from medical claims | 1 indicates incidence of at least one claim associated with co-morbidity flag since 2019 and 0 indicates its absence. | 32 |
| ICD-10-CM level 2 codes | Medical claim | ICD-10-CM level 2 codes (excluding those used as co-morbidity flags) occurring in at least one medical claim since 2019 are indicated by 1 and their absence by 0. | 1,018 |
| Medications | Pharmacy claim | 1 indicates incidence of at least one claim associated with the therapeutic class of medication since 2019 and 0 indicates its absence. | 282 |
| Age | Member information | Values above 0.5 SD were assigned 1 and those below 0.5 SD were assigned 0. Values in between were considered missing and assigned the value 3. | 1 |
| Risk scores based on Co-morbidity index | | Same as age | 4 |
| Lab analytes | Lab data | Same as age | 5 |

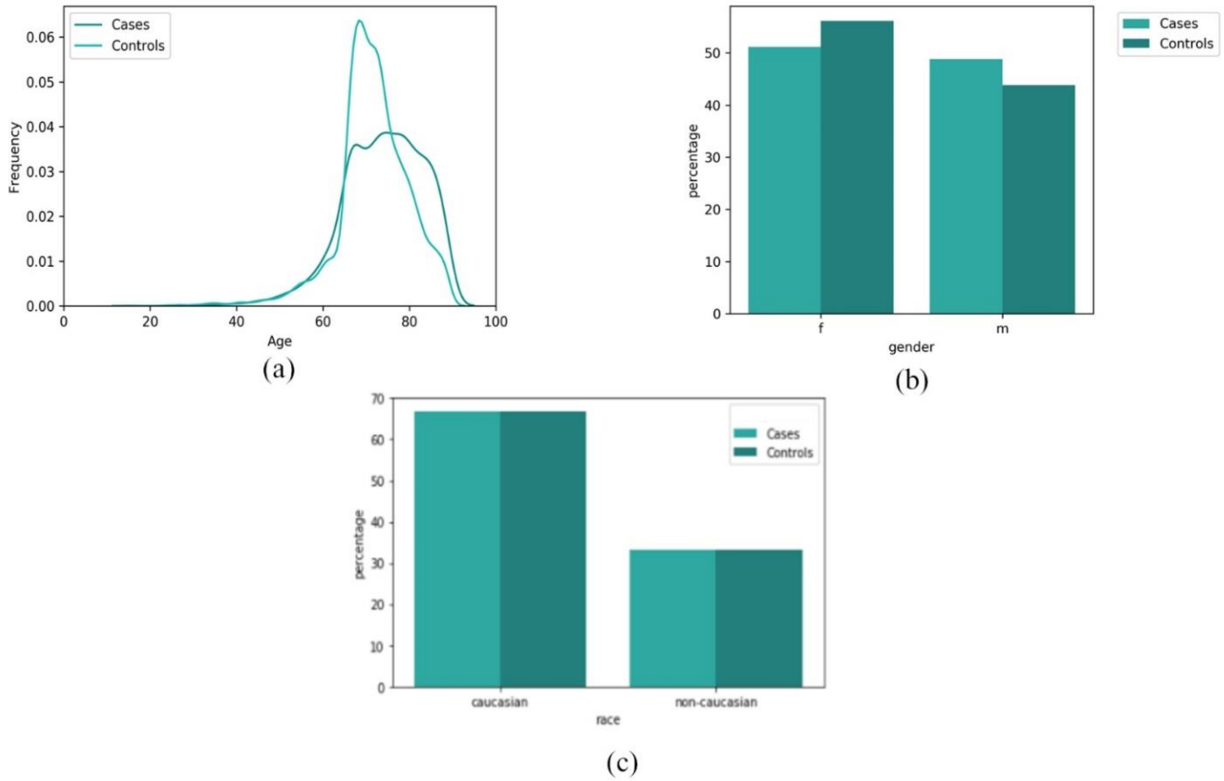


**Supplementary Figure 2.** Distribution of (a) age, (b) gender and (c) race for the full hospitalization risk cohort (Cohort 1, n=9,493 comprising of 3,183 cases and 6,310 controls).

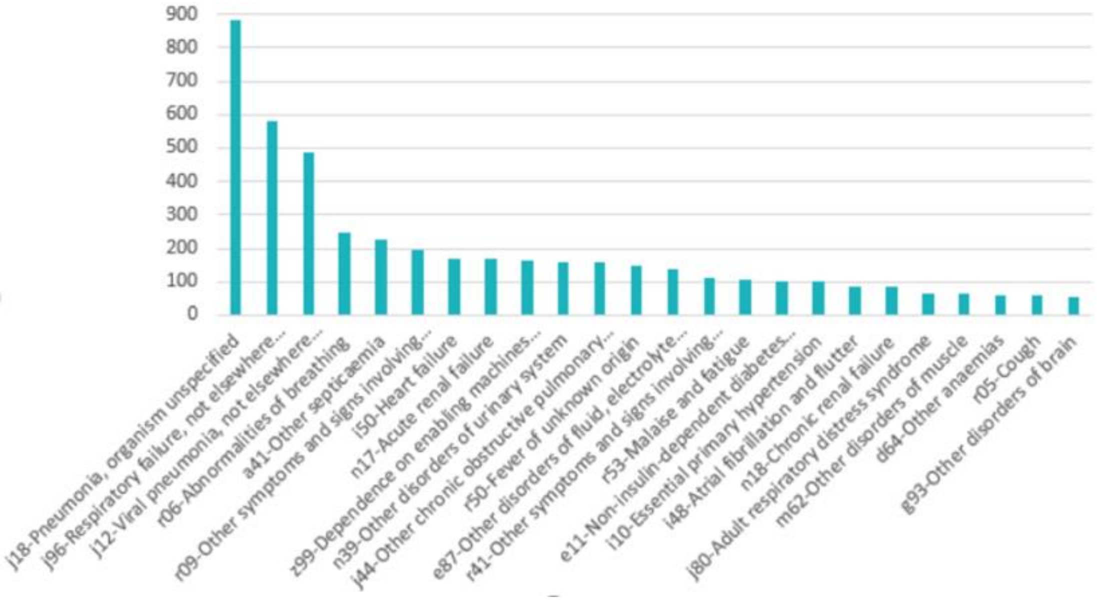


**Supplementary Figure 3.** Most frequently associated diagnoses associated with COVID-19 patients during their hospitalization in the full hospitalization risk cohort (Cohort 1).

**Supplementary Table 3.** Summary of combinatorial analysis results for the Hospitalization risk studies.

| | **Cohort 1** | **Cohort 2** |
|---|---|---|
| **Cases** | 3,183 (males: 1,549, females: 1,634) | 581 (males: 295, females: 286) |
| **Controls** | 6,310 (males: 2,758, females: 3,538) | 1,000 (males: 438, females: 560) |
| **Features** | 1,339 | 1,344 |
| **False Discovery Rate (FDR)** | 0.01 | 0.01 |
| **Random Permutations** | 2,500 | 2,500 |
| **Disease Signatures (N-states)** | 1,147 | 32,242 |
| **Penetrance** | 67.29% | 82.61% |
| **Filtered Disease Signatures** | 255 | 531 |
| **Penetrance of Filtered Disease Signatures** | 54.97% | 59.72% |
| **Critical features** | 166 | 41 |

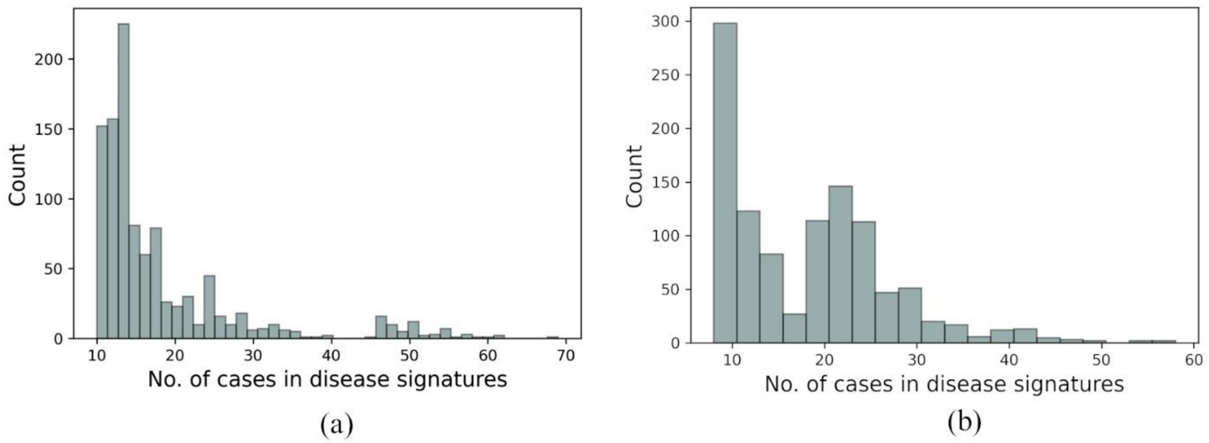


**Supplementary Figure 4.** Distribution of cases in COVID-19 disease signatures (excluding signatures that had any feature indicating absence of a disease diagnosis, symptom or medication) identified in (a) Cohort 1) and (b) Cohort 2.

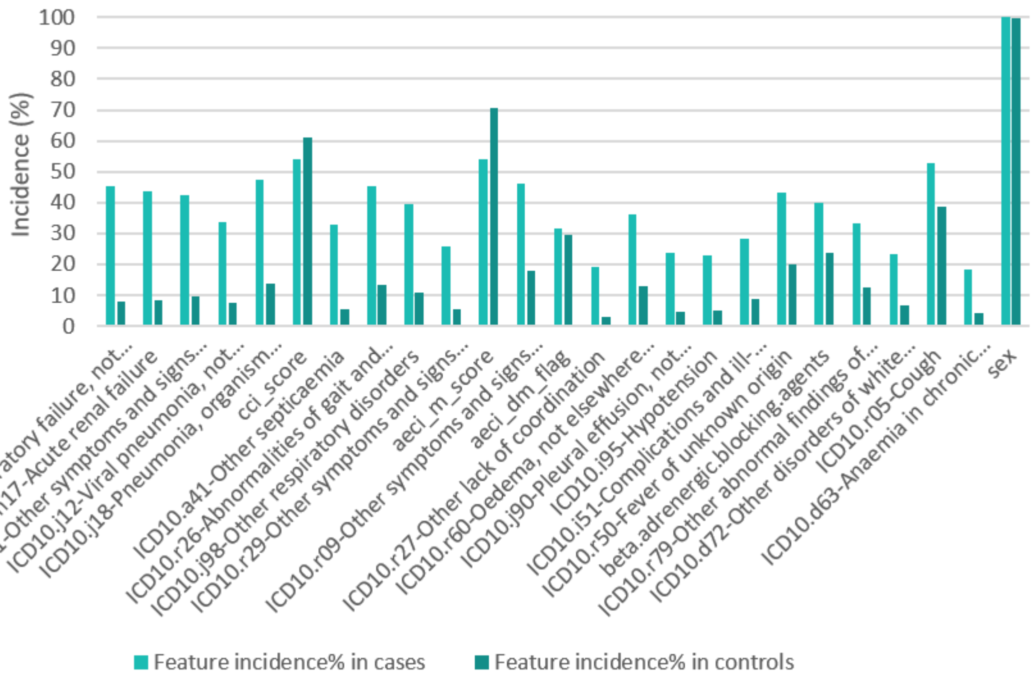


(a)

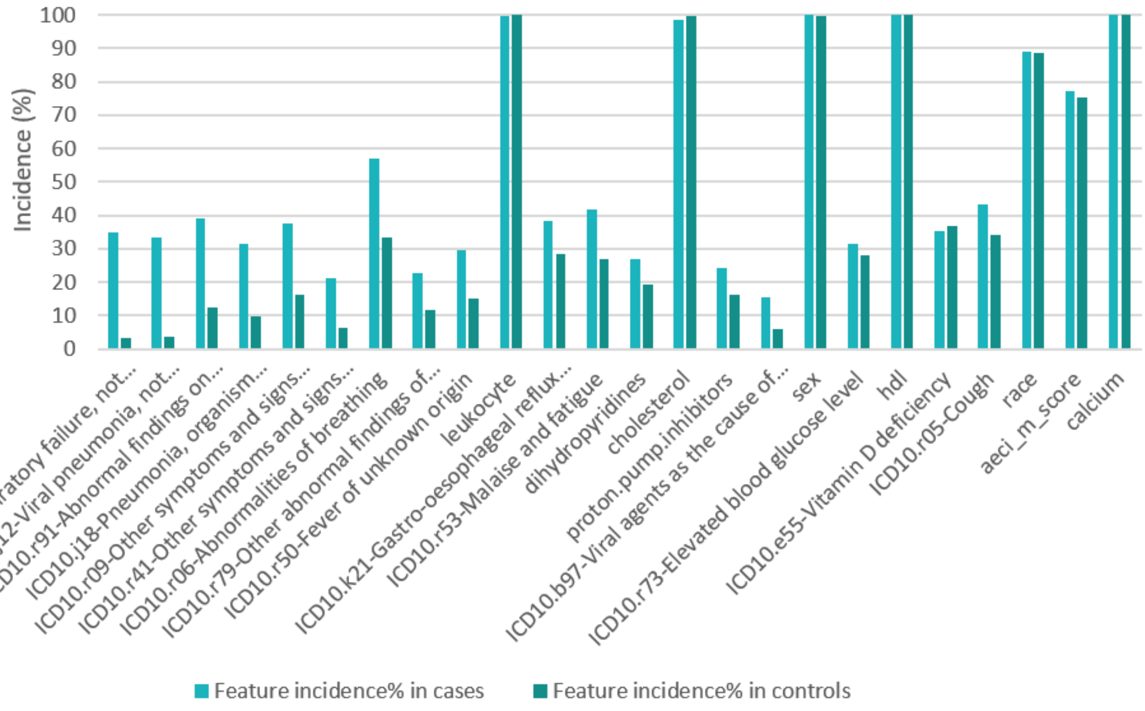


(b)

**Supplementary Figure 5.** Incidence of the top Random Forest **(**RF) scored critical features in cases and controls for (a) Cohort 1 and (b) Cohort 2.

**Supplementary Table 4.** Incidence of features derived from laboratory test results of patients in the cohort for the Hospitalized risk study using lab data. *p-values* were calculated to assess the association of each feature in the two COVID-19 hospitalized risk cohorts using two-sided Fisher's exact tests.

| **Lab analyte (normal range)** | **Lab analyte feature** | **Lab feature definition** | **Cases (n=581)** | **Controls (n=1000)** | **Two-sided p-value** | **One-sided p-value (lesser)** | **Disease signatures** |
|---|---|---|---|---|---|---|---|
| Calcium (8.6-10 mg/dl) | Calcium:0 | < 9.26 mg/dl in serum plasma | 289 | 770 | 0.0047 | 0.0024 | 18 |
| | Calcium:1 | > 9.71 mg/dl in serum plasma | 231 | 823 | 0.0047 | 0.998 | 4 |
| LDL (100 -129 mg/dl) | LDL:0 | < 78.23 mg/dl in serum plasma | 345 | 858 | 4.5e-09 | 2.8e-09 | 26 |
| | LDL:1 | >114.37 mg/dl in serum plasma | 224 | 984 | 4.5e-09 | 1 | 0 |
| HDL (40-60 mg/dl) | HDL:0 | < 44.35 mg/dl in serum plasma | 386 | 971 | 8.4e-10 | 4.2e-10 | 15 |
| | HDL:1 | > 66.16 mg/dl in serum plasma | 188 | 871 | 8.4e-10 | 1 | 4 |
| Triglycerides (100-150 mg/dl) | Triglycer:0 | < 73.05 mg/dl in serum plasma | 122 | 553 | 0.0002 | 0.9999 | 6 |
| | Triglycer:1 | > 206.20 mg/dl in serum plasma | 150 | 404 | 0.0002 | 0.00105 | 0 |
| Leukocyte count (5-10 k/mm$^3$) | Leukocyte:0 | < 5.28 k/mm3 in blood | 194 | 831 | 9.2e-09 | 3.9e-89 | 8 |
| | Leukocyte:1 | > 8.17 k/mm3 in blood | 240 | 542 | 9.2e-09 | 4.79e-07 | 9 |

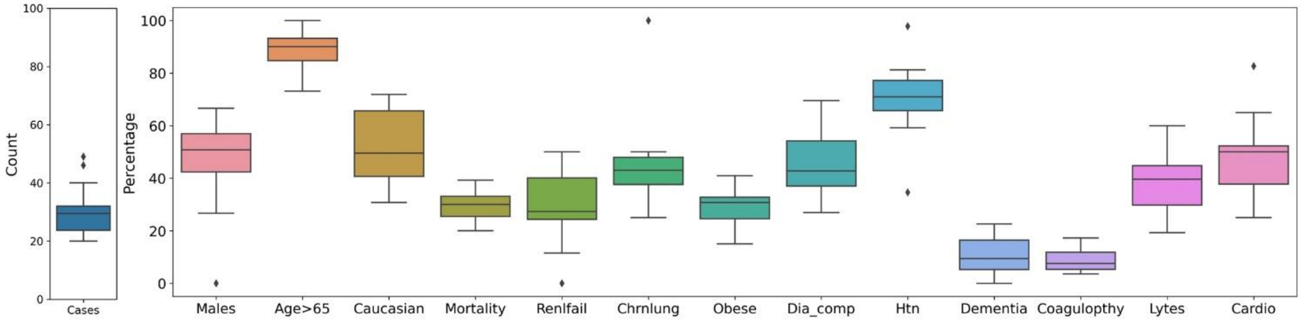


**Supplementary Figure 6.** Clinical characteristics of the disease signatures of hospitalized patients in Cohort 1 who had developed ARDS. Each boxplot captures the distribution of the incidence of a clinical feature in all ARDS disease signatures.

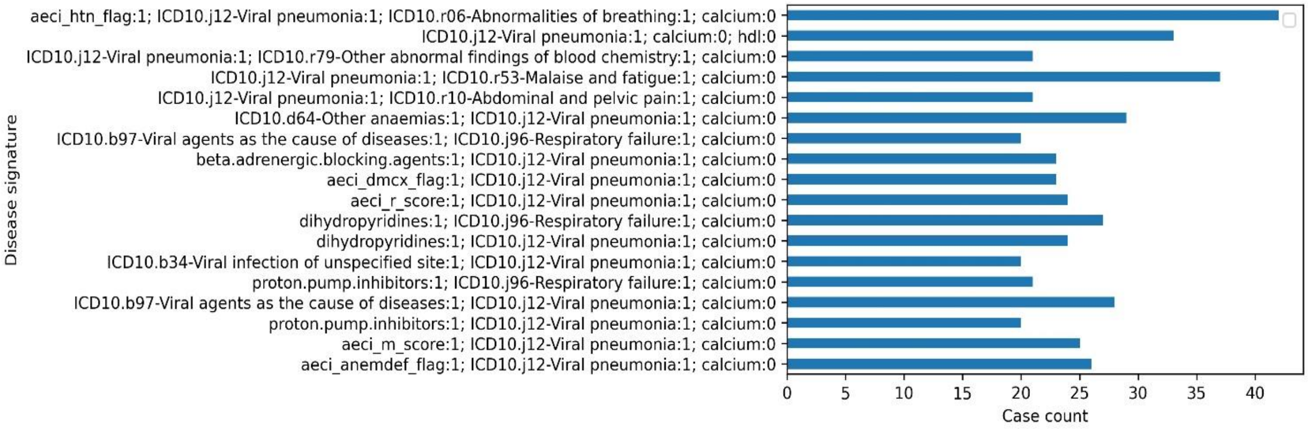


**Supplementary Figure 7.** Disease signatures with serum calcium levels below 9.11 mg/dl reported in Cohort 2.

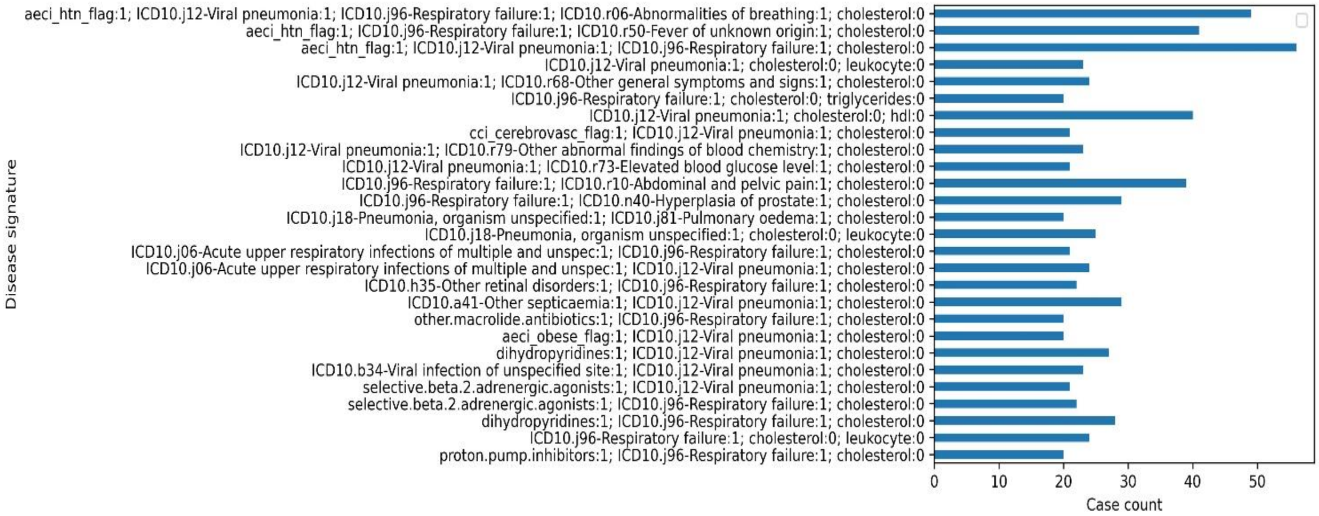


**Supplementary Figure 8.** Disease signatures with serum cholesterol (LDL) levels below 78.23 mg/dl reported in Cohort 2.

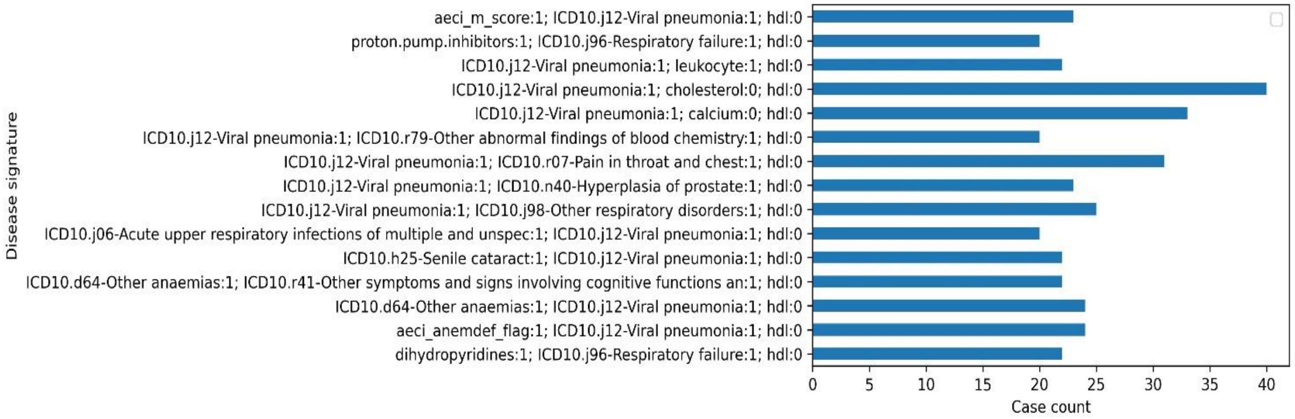

**Supplementary Figure 9.** Disease signatures with serum HDL levels below 44.35 mg/dl reported in Cohort 2.

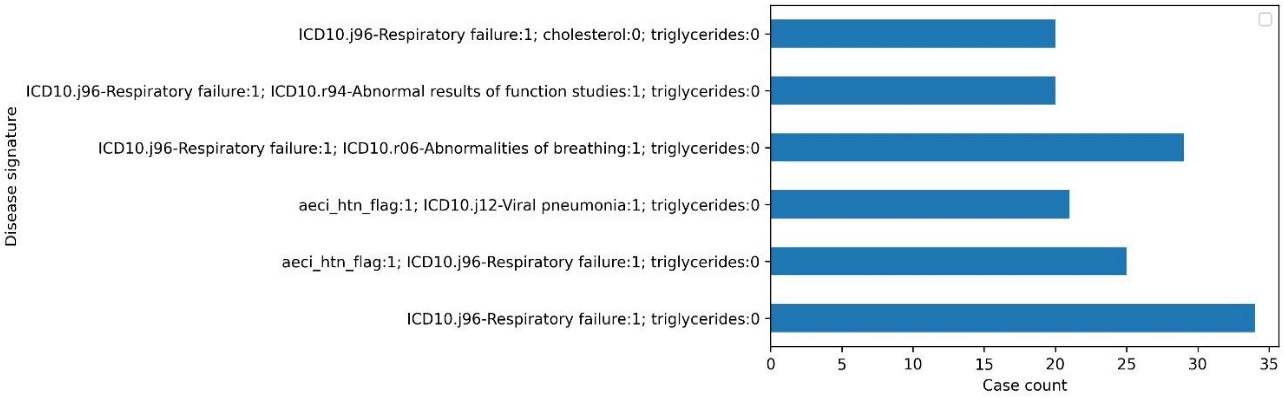


**Supplementary Figure 10.** Disease signatures with serum triglyceride levels below 73.05 mg/dl reported in Cohort 2.
